# Characteristics and Correlates of Older Smokers’ Experiences with E-Cigarette-Related Content on Social Media: Findings from a U.S.-Based Survey

**DOI:** 10.64898/2026.04.07.26350354

**Authors:** Ryan Dycus

**Affiliations:** Cornell Research on Tobacco Regulation

**Keywords:** Age, E-cigarettes, Harm reduction, Health communication, Older smokers, Risk perception, Social media, Tobacco harm reduction, Vaping

## Abstract

**Background:** Despite their potential to serve as a reduced-harm alternative to combustible tobacco, e-cigarette take-up remains low among older (45+) adult smokers, especially in the U.S. While social media is a known driver of vaping attitudes and behaviors in younger populations, its influence on older smokers is poorly understood. This paper provides the first focused analysis of e-cigarette-related social media exposure in this population, documenting its prevalence, characteristics, and attitudinal correlates.

**Methods:** Data come from an opt-in survey of U.S. adults (*N* = 974) recruited via Prolific, comprising three groups: (i) non-vaping smokers aged 45+ (*N* = 484), (ii) former-smoking vapers aged 45+ (*N* = 149), and (iii) any-vaping-status smokers aged 18-35 (*N* = 341). Descriptive statistics, weighted to U.S. population benchmarks, characterize self-reported exposure to e-cigarette-related content on social media. Logistic regressions estimate associations between exposure and intentions for future e-cigarette use, e-cigarette harm perceptions, and related attitudes.

**Results:** Older smokers (35.3%) reported exposure to e-cigarette-related content on social media less frequently than both older vapers (44.0%) and younger smokers (72.0%). For older smokers, e-cigarette health risks were the most frequently reported topic of content viewed, followed by youth vaping and e-cigarette addiction. Among this group, exposure was positively associated with stated intentions for future e-cigarette use. Exposure was not significantly associated with perceived e-cigarette harms for any group.

**Conclusions:** Findings provide suggestive evidence that social media exposure may promote e-cigarette adoption among older smokers. However, the cross-sectional design limits causal inference, and the observed associations may reflect selection bias or reverse causality. If a causal relationship exists, the patterns observed suggest that exposure influences e-cigarette adoption through mechanisms other than updating beliefs about e-cigarette risks. While these results tentatively support the potential of social media as a channel for older-smoker harm reduction, any policy applications must carefully weigh privacy concerns and risks to youth. Rigorous experimental studies are needed to confirm these findings and clarify how social media might be leveraged to improve public health outcomes among older smokers.

## 1 Introduction

Social media is replete with content discussing and depicting e-cigarettes and vaping. Millions of posts, videos, and images involving e-cigarettes scatter the various platforms that millions of Americans use daily [1–3]. This reality positions social media to serve as a critical venue in which attitudes, preferences, and beliefs regarding e-cigarettes are formed and modified. Consequently, social media’s coverage of e-cigarettes may influence behavior about their use, and thereby the state of public health. Examining the influence of e-cigarette-related content on social media is therefore important for public health, particularly with regard to key populations such as older adult smokers.

A growing, youth-oriented literature assesses both the supply and consumption of e-cigarette-related social media content with an orientation toward its implications for nicotine product use. With respect to the former, many content analyses document and measure the valence [4–7], themes [2, 6, 8–11], and informational validity [1] of e-cigarette social media content across various platforms. However, such analyses focus on the overall pool of content available rather than on user-specific exposure, and are inherently limited by the algorithmically curated, personalized nature of social media feeds, preventing clear inference about who sees what. Characterization analyses build upon this foundation by shifting attention to the viewer’s perspective, using survey data to describe the nature of content individuals report encountering in their own feeds [12, 13]. In addition to characterization analyses, the consumption-side of research also includes associational analyses of survey data [14–16] and experimental work [17–22], which seek to directly assess how exposure to e-cigarette-related content shapes individuals’ attitudes and behaviors regarding e-cigarettes and other nicotine products.

Nevertheless, the youth focus of existing work leaves a substantial gap in the literature with regard to the evaluation of older tobacco users, particularly smokers. To the best of the author’s knowledge, only four published studies pertaining to e-cigarette-related social media exposure analyze data that include individuals aged 45 and older, and the analyses related to social media in these studies are dated and not focused on older populations. Emery et al. and Emory et al. [23, 24], which use the same data, and Bauhoff et al. [25] provide summary measures of e-cigarette content exposure within their respective samples, and the former two also report on content engagement (e.g., whether sampled individuals had shared e-cigarette-related posts). Majmundar et al. [26] assess the link between what they define as e-cigarette-related social media “engagement” (including simple exposure) and support for stricter e-cigarette regulation, finding a negative association. Although these studies established social media’s importance as an information vector for e-cigarettes, their respective data are at least eight years old, limiting their current relevance in the especially tumultuous e-cigarette market. Moreover, none of these studies provide heterogeneity analyses by age in general or by age and tobacco use status in particular—a limitation that is especially consequential given the massive public health implications of later-in-life smoking.

Older (45+) smokers play a central role in the ongoing burden of tobacco-related disease in the United States. Although smoking prevalence declined substantially between 2011 and 2022, the decline was markedly smaller among adults aged 40–64 compared with younger cohorts, and the smoking rate among adults aged 65+ showed no decrease at all during this period [27]. As a result, older adults now comprise the majority of the smoking population. According to the 2023 National Health Interview Survey (NHIS), adults aged 45+ accounted for 58.6% of current smokers and 62.3% of everyday smokers [28]. Overall, 11.8% of the U.S. population aged 45+ and 8.5% of those aged 65+ were current smokers in 2023, amounting to 16.7 million and 2.3 million people, respectively [28, 29]. Promoting quitting among older smokers remains an essential public health priority. While estimates range according to statistical methodology and data, previous studies estimate that smokers can gain back between three to six years of life expectancy by quitting at age 50 [30, 31], with measurable gains still possible as late as 75 [32]. Beyond mortality, those who quit later in adulthood also experience improvements in quality of life, such as better respiratory function [33]. Given the size of the problem posed by later-in-life smoking and the immense potential benefits that accrue from its reduction, it is critical to consider all viable cessation and harm-reduction tools available to this population—including, increasingly, e-cigarettes.

On account of their relative safety and cessation efficacy, e-cigarettes hold considerable potential for smoking harm reduction, yet are presently underutilized by older smokers. Both public agencies [34, 35] and scientific experts [36] deem e-cigarettes considerably safer than combustible tobacco. This consensus position is reflected in the official policy of the U.S. Food and Drug Administration, whose *Relative Risks of Tobacco Products* guidance statement advises that smokers can improve their health by completely switching to e-cigarettes [37]. Evidence also supports their cessation efficacy: a Cochrane meta-analysis found e-cigarettes to be the most effective clinical intervention for promoting quitting [38], and findings from a recent naturalistic RCT suggest these benefits extend beyond clinical settings [39]. Despite this potential, e-cigarette take-up among older smokers in the U.S. remains low. According to recent data from the Wave 7 (2022) Population Assessment of Tobacco and Health, only 5.2% of smokers over 45 who attempted to quit reported trying e-cigarettes as a cessation aid (compared to 11.3% for those 18–44). Additionally, only 1.2% of non-vaping smokers over 45 in Wave 6 (2021) transitioned to being non-smoking vapers in Wave 7 (compared to 5.4% for those 18–44) [40]. These statistics underscore the need to better understand the factors that reinforce or reduce older smokers’ reluctance to adopt e-cigarettes, such as the influence of e-cigarette-related social media content exposure.

This descriptive, survey-based study addresses this knowledge gap by providing an initial assessment of the nexus between social media and older smokers’ attitudes and behaviors regarding e-cigarettes. The study analyzes responses from three sampling groups: (i) 484 non-vaping current smokers aged 45 and older; (ii) 149 non-smoking current vapers aged 45 and older; and (iii) 341 current smokers aged 18–35 of varying vaping statuses. While the analysis focuses on the first group, the latter two serve as points of comparison along the dimensions of age and tobacco-use status. The analysis proceeds with two aims. First, it conducts a comparative characterization analysis, in which the nature of participants’ e-cigarette-related social media exposure is described and contrasted across the three sampling groups. In this respect, the study parallels Vogel et al. [13], which provides a comparative characterization analysis by nicotine product use status for a sample of U.S. adolescents and young adults. Second, it presents associational analyses that examine how e-cigarette-related social media exposure covaries with attitudes and preferences toward e-cigarettes among participants. In doing so, the study extends the types of associational analyses reviewed in Donaldson et al. [15] and Rutherford et al. [16] to understudied populations.

## 2 Data and Methods

### 2.1 Survey Procedure and Sample

The survey data for this study were collected as part of a larger pilot from an opt-in sample on Prolific: a dedicated participant platform that provides high-quality human subject data for academic research [41, 42], with prior use in tobacco research [43]. Three sampling groups were recruited using Prolific’s prescreening feature: (1) non-vaping, current smokers aged 45 and older (*over-45 exclusive smokers*); (2) current vaping, former smokers aged 45 and older (*over-45 exclusive vapers*); and (3) current smokers of any vaping status aged 35 and younger (*under-36 smokers*). All participants were English proficient, U.S. residents. Definitions of nicotine product use status correspond to Prolific’s built-in prescreening criteria. Specifically, current smokers are defined as those who smoke at least five cigarettes daily for at least a year; current vapers as those who vape at least once daily; former smokers as those who previously smoked at least five cigarettes daily for a year but no longer smoke; and non-vapers as those who have vaped fewer than 20 times in their lifetime. It should be emphasized that the “current user” definitions here differ from standard definitions used in public health surveillance, which typically define current smokers or vapers either as individuals who report having used the product at least once in the past 30 days, or who report using them “some days” or “every day” rather than “not at all” [28, 44].

A total of 980 respondents completed the survey, of which six were excluded due to missing key demographic data. This resulted in a final analytic sample of 484 over-45 exclusive smokers, 149 over-45 exclusive vapers, and 341 under-36 smokers, for a total sample size of *N* = 974. Data were collected in November, 2024. While a preregistration plan was submitted to OSF for the broader pilot (link), this plan did not outline the specific analysis conducted in this article; the results presented should therefore be considered exploratory.

Summary details about the demographic composition of the sample are presented in Table 1, which reports mean values of various demographic and tobacco use characteristics by sampling group. Broadly speaking, the sampling groups are not representative of their respective national counterparts in terms of demographics (e.g., the percentage of sampled older smokers who are female (72.7%) is higher than recent national estimates, which hover around 50% [28, 44]). At the same time, several patterns of tobacco use within the sample are roughly consistent with national statistics (e.g., older smokers smoke more cigarettes per day than young smokers [44]). All analyses account for demographic imbalances through weighting or covariate adjustment.

**Table 1:**
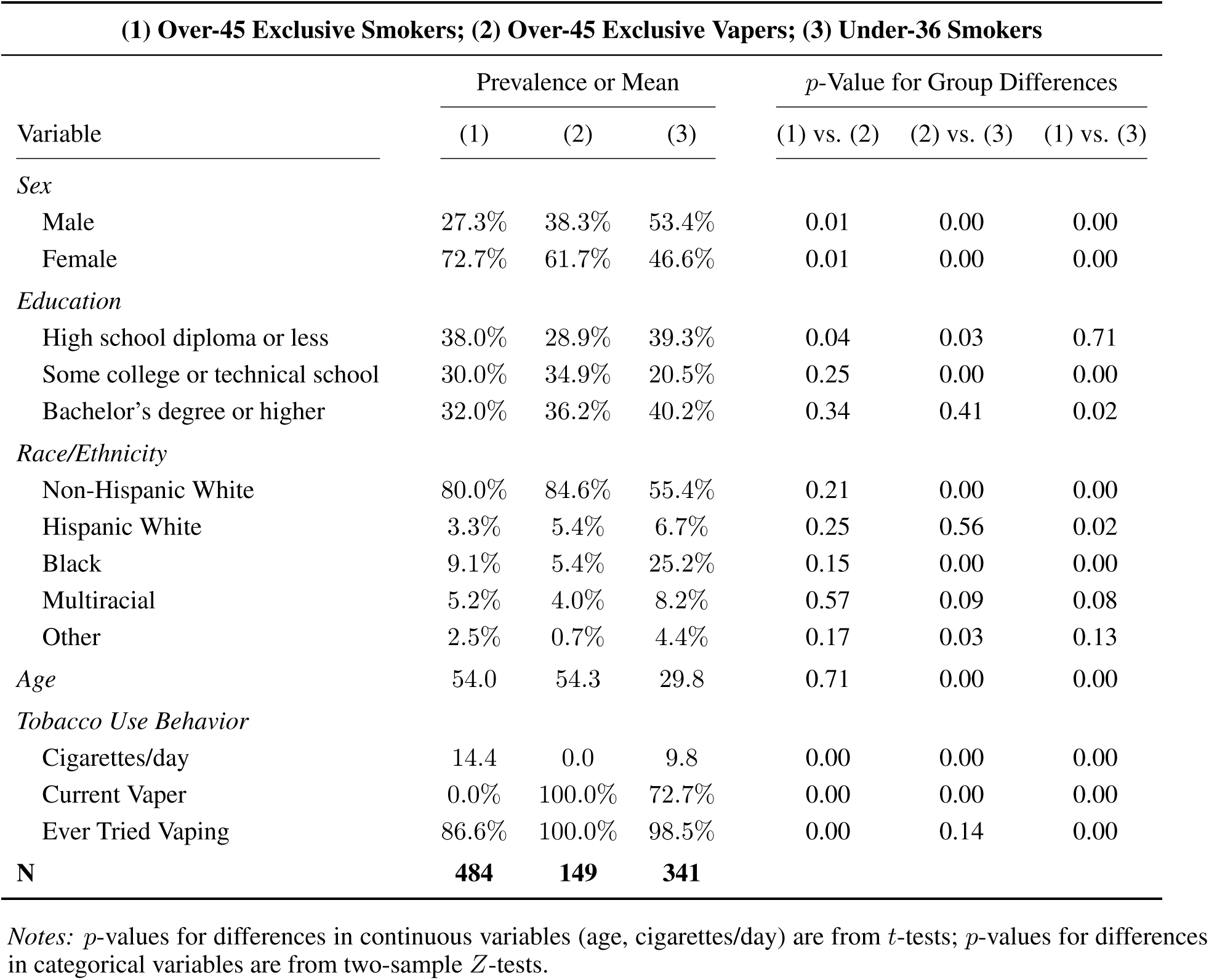
Sample Descriptives.

| (1) Over-45 Exclusive Smokers; (2) Over-45 Exclusive Vapers; (3) Under-36 Smokers |  |  |  |  |  |  |
| --- | --- | --- | --- | --- | --- | --- |
| Variable | Prevalence or Mean |  |  | *p*-Value for Group Differences |  |  |
|  | (1) | (2) | (3) | (1) vs. (2) | (2) vs. (3) | (1) vs. (3) |
| *Sex* |  |  |  |  |  |  |
| Male | 27.3% | 38.3% | 53.4% | 0.01 | 0.00 | 0.00 |
| Female | 72.7% | 61.7% | 46.6% | 0.01 | 0.00 | 0.00 |
| *Education* |  |  |  |  |  |  |
| High school diploma or less | 38.0% | 28.9% | 39.3% | 0.04 | 0.03 | 0.71 |
| Some college or technical school | 30.0% | 34.9% | 20.5% | 0.25 | 0.00 | 0.00 |
| Bachelor's degree or higher | 32.0% | 36.2% | 40.2% | 0.34 | 0.41 | 0.02 |
| *Race/Ethnicity* |  |  |  |  |  |  |
| Non-Hispanic White | 80.0% | 84.6% | 55.4% | 0.21 | 0.00 | 0.00 |
| Hispanic White | 3.3% | 5.4% | 6.7% | 0.25 | 0.56 | 0.02 |
| Black | 9.1% | 5.4% | 25.2% | 0.15 | 0.00 | 0.00 |
| Multiracial | 5.2% | 4.0% | 8.2% | 0.57 | 0.09 | 0.08 |
| Other | 2.5% | 0.7% | 4.4% | 0.17 | 0.03 | 0.13 |
| *Age* | 54.0 | 54.3 | 29.8 | 0.71 | 0.00 | 0.00 |
| *Tobacco Use Behavior* |  |  |  |  |  |  |
| Cigarettes/day | 14.4 | 0.0 | 9.8 | 0.00 | 0.00 | 0.00 |
| Current Vaper | 0.0% | 100.0% | 72.7% | 0.00 | 0.00 | 0.00 |
| Ever Tried Vaping | 86.6% | 100.0% | 98.5% | 0.00 | 0.14 | 0.00 |
| **N** | **484** | **149** | **341** |  |  |  |

*Notes:* *p*-values for differences in continuous variables (age, cigarettes/day) are from *t*-tests; *p*-values for differences in categorical variables are from two-sample *Z*-tests.

### 2.2 Survey Measures

Participants were asked a number of questions about their attitudes and experiences pertaining to e-cigarettes, including their experiences with e-cigarette-related content on social media. Additionally, over-45 exclusive smokers were asked about their intentions for future e-cigarette use. All survey items considered in this study permitted respondents to opt out of providing prespecified responses. Please see Appendix A for exact wordings of all items.

#### 2.2.1 General Social Media Use

All participants were asked “*Do you regularly use social media?*,” with response options *yes* or *no*. All who answered *yes* were then asked two additional questions about which platforms they used and how frequently they used social media in general. Respondents indicated their use of specific platforms by selecting any that applied from *Facebook*, *Instagram*, *TikTok*, *X/Twitter*, *YouTube*, or *Other*. They indicated their use frequency by selecting from a six-item list ranging from *multiple times a day* to *about once a month*.

#### 2.2.2 E-Cigarette-Related Social Media Exposure

All participants, including those who did not indicate regular social media use, were asked whether they had “*ever viewed any content (posts, pictures, videos, memes, etc.) on social media discussing, depicting, or mentioning e-cigarettes or their use?*,” with response options *yes* or *no*. Respondents who answered *yes* were then asked further questions about their experiences with e-cigarette-related content, including the topics of content viewed, the platforms on which viewing occurred, whether the content was searched for or recommended, how recently content was viewed, and how many times they recalled viewing.

Participants were asked to indicate all topics of e-cigarette-related content they had “noticed,” as well as the single topic they had “noticed the most.” The response options—drawn from prior content analyses [1, 2, 6, 8–11]—were the same for both: *health risks*; *health benefits for smokers*; *industry news*; *youth e-cigarette use*; *addiction*; *user guides*; *product reviews*; *lifestyle content portraying e-cigarettes as cool, sexy, or fashionable*; and *other*. A minority of respondents (14.7%) selected a “most noticed” topic that they had not listed among the “any noticed” topics. To correct for this inconsistency, “most noticed” topics were imputed as additions to the “any noticed” lists. This correction is supported by prior studies that demonstrate a systematic tendency for participants to underreport when answering multiple-response questions [45, 46].

To provide concise summaries of the overall slant of e-cigarette-related content seen by respondents, reported topics were grouped by their likely persuasive valence—that is, the extent to which they are typically framed in a way that promotes or discourages vaping. Prior literature suggests that certain topics and themes largely align with a particular valence orientation. Specifically, *product reviews*, *user guides*, *health benefits for smokers*, and *lifestyle content* were categorized as positively valenced [6, 9–11]; *health risks*, *addiction*, and *youth vaping* were categorized as negatively valenced [1, 2, 8]. The responses *other* and *industry news* were excluded from this scheme due to a lack of empirical evidence supporting a consistent persuasive framing. Using these categorizations, three mutually exclusive indicator variables were constructed from the “topics noticed” lists: (i) exposure to at least one positive and no negative topics (only positive topics seen), (ii) exposure to at least one negative and no positive topics (only negative topics seen), and (iii) exposure to both positive and negative topics.

The item that elicited the platforms on which e-cigarette-related content was seen had the same response options as that for general social media use. The extent to which participants searched for or were recommended e-cigarette content was measured on a five-point scale ranging from *none of the content was recommended to me* to *all of the content was recommended to me*. Finally, participants reported their viewing recency on a six-point scale ranging from *within the past week* to *more than two years ago*, and their total e-cigarette content encounters on a six-point scale ranging from *less than 10 times* to *more than 200 times*.

#### 2.2.3 Other E-Cigarette Measures

Over-45 exclusive smokers were asked about their intentions for future e-cigarette use through three questions: (i) their general interest in using e-cigarettes in the future (four-point response scale from *not interested at all* to *extremely interested*); (ii) how likely they would be to consider switching to e-cigarettes if “*cigarette prices were to increase by $4 per pack*” (five-point response scale from *extremely unlikely* to *extremely likely*); and (iii) how likely they would be to consider switching if they “*did not have to pay for e-cigarettes*” (five-point response scale from *extremely unlikely* to *extremely likely*).

All participants were asked three questions pertaining to the potential harms of e-cigarettes: (i) how they viewed the general harmfulness of e-cigarettes for health (four-point response scale from *not harmful at all* to *extremely harmful*); (ii) how they compared the health harms of e-cigarettes to cigarettes (five-point response scale from “e-cigarettes are…” *much less harmful to health* to *much more harmful to health*); and (iii) how they compared the addictiveness of e-cigarettes to cigarettes (five-point response scale from “e-cigarettes are…” *much less addictive* to *much more addictive*).

Finally, all participants answered one item each on e-cigarette regulation, legality, and marketing intentions. Each was framed as a statement, with responses on a five-point scale ranging from *strongly disagree* to *strongly agree*: (i) “*the government should spend money on public service announcements aimed at discouraging youth from using e-cigarettes*”; (ii) “*a lot of e-cigarette sales today are illicit*”; and (iii) “*e-cigarettes are made with adult smokers in mind*.”

### 2.3 Statistical Analysis

This study has two analytical aims. The first is to characterize the sample’s e-cigarette-related social media exposure and estimate how the nature of this exposure varies across the study’s three sampling groups: over-45 exclusive smokers, over-45 exclusive vapers, and under-36 smokers. The second aim is to estimate within-group associations between exposure and e-cigarette beliefs, attitudes, and (for older smokers) intentions for future e-cigarette use.

#### 2.3.1 Comparative Characterization Analysis

Responses to social media items—both general and e-cigarette-specific—were summarized by sampling group using post-stratification-weighted prevalences (reports of raw prevalences can be found in Appendix B). Specifically, post-stratification weights were applied within each sampling group to align respondents to the demographic distribution of their corresponding subgroup in the 2023 NHIS along three dimensions: sex (male vs. female), educational attainment (bachelor’s degree or higher vs. lower), and race/ethnicity (non-Hispanic white vs. all other groups). Standard errors for the weighted prevalences were obtained using Taylor series linearization. Significant differences in response frequencies across sampling groups were assessed using design-adjusted Wald tests. Each option in multiple-response items was analyzed as a separate binary indicator, coded 1 if selected. For the item regarding past exposure to e-cigarette-related social media content, opt-out responses were retained and reported, given their high number and the item’s centrality to the study; for all other items, opt-outs were dropped. Response options to single-selection ordinal items with sparse categories were collapsed into adjacent categories to avoid unstable estimates.

#### 2.3.2 Associational Analysis

Logistic regression was used to assess sampling group-specific associations between exposure to e-cigarette-related content and survey measures of e-cigarette beliefs, attitudes, and (for over-45 exclusive smokers only) intentions for future e-cigarette use. Weighted by-group summaries of responses to these measures are presented in Appendix C. Content exposure was modeled as a binary regressor, coded 1 if the participant answered *yes* to the exposure question and 0 if they answered *no* or opted out by selecting “I don’t recall.” Opt-outs were retained to preserve statistical power; however, because they may represent a substantively different group than respondents who answered *no*, a sensitivity analysis excluding them was conducted (Appendix D), yielding directionally similar results. Ordered categorical outcomes were dichotomized at single cutoffs selected to balance interpretability, stability, and consistency; cutoff codings are reported in Table 4. Robustness analyses using ordinal logit models and alternative binary cutoffs are presented in Appendix D. Associations for each sampling group were obtained from exposure-by-group interaction terms. All associational models controlled for sex, education, and race/ethnicity using the levels reported in Table 1. Primary estimates were unweighted, consistent with recommendations that regression models including weight-defining covariates can recover consistent conditional associations with improved statistical efficiency [47]. As a supporting diagnostic, post-stratification weighted estimates (Appendix D) yielded qualitatively similar point estimates with generally larger confidence intervals. Standard errors were estimated with Huber-White robust estimators, and statistical significance was assessed using Wald tests. Results are presented on the odds ratio (OR) scale with 95% confidence intervals.

## 3 Results

Table 2 presents post-stratification–weighted estimates of general social media use by sampling group. Regular use of social media was reported by 91.4% of over-45 exclusive smokers, 93.6% of over-45 exclusive vapers, and 95.1% of under-36 smokers. Most respondents in each group reported using social media multiple times per day. Facebook and YouTube were the two most commonly used platforms across all groups. Among older smokers (80.7%) and older vapers (81.2%), Facebook was the most frequently reported, whereas YouTube was most frequently reported among younger smokers (89.1%). Consistent with findings from the representative survey analysis by Gottfried [3], the two older groups were significantly less likely than younger smokers to report using Instagram and TikTok (*p < .*01 for all comparisons).

**Table 2:** Characteristics of General Social Media Use.

| **(1) Over-45 Exclusive Smokers; (2) Over-45 Exclusive Vapers; (3) Under-36 Smokers** |  |  |  |  |  |  |
| --- | --- | --- | --- | --- | --- | --- |
| Survey Measure | Weighted Prevalence (95% CI) |  |  | *p*-Value for Group Differences |  |  |
|  | (1) | (2) | (3) | (1) vs. (2) | (2) vs. (3) | (1) vs. (3) |
| *Use Social Media Regularly?* |  |  |  |  |  |  |
| Yes | 91.4% (88.1, 94.7) | 93.6% (89.2, 98.0) | 95.1% (92.3, 98.0) | 0.43 | 0.57 | 0.10 |
| No | 8.6% (5.3, 11.9) | 6.4% (2.0, 10.8) | 4.9% (2.0, 7.7) | 0.43 | 0.57 | 0.10 |
| **N** | **483** | **149** | **338** |  |  |  |
| *Social Media Use Frequency* |  |  |  |  |  |  |
| Less often than daily | 5.7% (2.8, 8.6) | 7.3% (2.7, 11.9) | 1.6% (0.1, 3.2) | 0.57 | 0.02 | 0.01 |
| Once per day | 16.9% (12.8, 21.0) | 19.4% (11.6, 27.2) | 9.9% (5.9, 13.9) | 0.59 | 0.03 | 0.02 |
| Multiple times per day | 77.3% (72.6, 82.1) | 73.3% (64.8, 81.8) | 88.5% (84.3, 92.6) | 0.42 | 0.00 | 0.00 |
| **N** | **443** | **140** | **321** |  |  |  |
| *Platforms Used* |  |  |  |  |  |  |
| Facebook | 80.7% (75.7, 85.6) | 81.2% (74.1, 88.4) | 82.8% (77.9, 87.8) | 0.90 | 0.72 | 0.54 |
| Instagram | 47.1% (41.2, 53.0) | 47.5% (38.1, 57.0) | 68.3% (62.1, 74.4) | 0.94 | 0.00 | 0.00 |
| TikTok | 40.1% (34.4, 45.8) | 34.8% (25.8, 43.7) | 53.3% (46.8, 59.8) | 0.33 | 0.00 | 0.00 |
| X/Twitter | 33.9% (28.3, 39.4) | 47.8% (38.3, 57.3) | 49.5% (43.0, 56.0) | 0.01 | 0.78 | 0.00 |
| YouTube | 79.9% (75.5, 84.3) | 78.7% (71.3, 86.0) | 89.1% (85.2, 92.9) | 0.77 | 0.01 | 0.00 |
| Other | 9.8% (6.4, 13.1) | 10.6% (5.1, 16.1) | 7.5% (4.0, 11.0) | 0.80 | 0.36 | 0.36 |
| **N** | **443** | **140** | **321** |  |  |  |

*Notes:* Item response prevalences are post-stratification-weighted within each sampling group to match the 2023 NHIS by sex, race/ethnicity, and educational attainment (see Section 2.3.1 for details). *p*-values for differences in weighted responses across sampling groups are from design-adjusted Wald tests. 95% confidence intervals are shown in parentheses.

Tables 3a and 3b present weighted estimates of e-cigarette-related social media exposure by sampling group. Over-45 exclusive smokers (35.3%) and over-45 exclusive vapers (44.0%) reported significantly lower exposure than under-36 smokers (72.0%), whose prevalence falls within the range of estimates for young adult and adolescent populations in recent nationally representative studies [12, 13]. The difference between older vapers and older smokers (8.7 percentage points) narrowly missed conventional thresholds for marginal significance (*p ≈ .*11), but the magnitude indicates a potentially meaningful disparity, especially in light of unweighted comparisons (see Appendix B). A notable minority of respondents were unable to recall whether they had been exposed to e-cigarette-related content—including 13.4% of older smokers, 11.0% of older vapers, and 5.6% of younger smokers. Among those reporting exposure, Facebook and YouTube were the most frequently indicated platforms across all groups. Among older smokers, 45.6% indicated exposure on Facebook and 52.7% on YouTube; corresponding figures were 49.4% and 46.1% for older vapers, and 57.6% and 53.4% for younger smokers. Exposure on TikTok and Instagram was significantly higher among younger smokers compared with both older groups (*p < .*01 for all four comparisons), consistent with cross-group differences in general platform use.

**Table 3a:** Characteristics of E-Cigarette-Related Social Media Exposure.

| **(1) Over-45 Exclusive Smokers; (2) Over-45 Exclusive Vapers; (3) Under-36 Smokers** |  |  |  |  |  |  |
| --- | --- | --- | --- | --- | --- | --- |
| Survey Measure | Weighted Prevalence (95% CI) |  |  | *p*-Value for Group Differences |  |  |
|  | (1) | (2) | (3) | (1) vs. (2) | (2) vs. (3) | (1) vs. (3) |
| *Ever seen e-cig social media content* |  |  |  |  |  |  |
| Yes | 35.3% (29.8, 40.7) | 44.0% (34.9, 53.1) | 72.0% (66.3, 77.7) | 0.11 | 0.00 | 0.00 |
| No | 51.3% (45.7, 57.0) | 45.0% (36.1, 54.0) | 22.4% (17.1, 27.7) | 0.24 | 0.00 | 0.00 |
| Don't recall | 13.4% (9.7, 17.1) | 11.0% (4.6, 17.3) | 5.6% (2.7, 8.5) | 0.52 | 0.13 | 0.00 |
| **N** | **484** | **149** | **341** |  |  |  |
| *Any topics of content noticed* |  |  |  |  |  |  |
| E-cig health risks | 64.4% (55.0, 73.9) | 58.7% (45.0, 72.5) | 63.9% (56.6, 71.1) | 0.50 | 0.52 | 0.92 |
| E-cig health benefits for smokers | 14.9% (8.4, 21.4) | 25.3% (13.6, 37.1) | 15.7% (10.5, 21.0) | 0.13 | 0.14 | 0.84 |
| Youth vaping | 53.9% (43.9, 63.8) | 29.7% (16.2, 43.1) | 55.0% (47.5, 62.5) | 0.00 | 0.00 | 0.85 |
| E-cig industry news | 13.6% (7.1, 20.1) | 26.9% (15.4, 38.5) | 24.4% (18.0, 30.8) | 0.05 | 0.71 | 0.02 |
| E-cig addiction | 42.2% (32.4, 51.9) | 27.0% (13.8, 40.2) | 49.7% (42.2, 57.3) | 0.07 | 0.00 | 0.23 |
| Guides on e-cig use | 12.9% (6.2, 19.6) | 27.2% (14.8, 39.6) | 14.4% (9.3, 19.6) | 0.05 | 0.06 | 0.72 |
| E-cig product reviews | 27.4% (18.8, 36.0) | 66.0% (52.2, 79.7) | 40.9% (33.5, 48.4) | 0.00 | 0.00 | 0.02 |
| Lifestyle content involving e-cigs | 30.7% (21.4, 39.9) | 18.7% (6.3, 31.1) | 36.2% (28.9, 43.4) | 0.13 | 0.02 | 0.36 |
| Other | 4.4% (0.2, 8.7) | 6.5% (0.6, 12.5) | 0.8% (0.0, 2.0) | 0.57 | 0.06 | 0.11 |
| Num. of topics indicated (mean) | 2.6 (2.4, 2.9) | 2.9 (2.5, 3.2) | 3.0 (2.8, 3.2) | 0.30 | 0.45 | 0.03 |
| **N** | **153** | **63** | **242** |  |  |  |
| *Derived topical valence measures* |  |  |  |  |  |  |
| Only positive topics seen | 19.8% (11.9, 27.7) | 30.1% (17.1, 43.0) | 12.7% (7.7, 17.7) | 0.18 | 0.01 | 0.14 |
| Only negative topics seen | 45.5% (35.4, 55.6) | 21.3% (10.5, 32.0) | 33.0% (25.9, 40.1) | 0.00 | 0.07 | 0.05 |
| Both positive and negative topics seen | 11.5% (7.9, 15.0) | 20.4% (12.3, 28.5) | 38.0% (31.8, 44.1) | 0.05 | 0.00 | 0.00 |
| **N** | **151** | **61** | **240** |  |  |  |
| *Topic of content noticed most* |  |  |  |  |  |  |
| E-cig health risks | 35.0% (25.5, 44.6) | 28.7% (15.1, 42.3) | 35.4% (28.3, 42.6) | 0.46 | 0.39 | 0.95 |
| E-cig health benefits for smokers | 1.3% (0.0, 3.0) | 2.9% (0.0, 7.0) | 1.5% (0.1, 2.9) | 0.47 | 0.52 | 0.87 |
| Youth vaping | 22.3% (13.6, 31.0) | 6.8% (0.4, 13.3) | 16.7% (10.8, 22.5) | 0.01 | 0.03 | 0.29 |
| E-cig industry news | 1.3% (0.0, 2.7) | 7.9% (0.8, 15.0) | 5.3% (2.0, 8.5) | 0.08 | 0.51 | 0.03 |
| E-cig addiction | 7.2% (2.2, 12.2) | 3.7% (0.0, 8.1) | 9.3% (5.1, 13.5) | 0.30 | 0.07 | 0.53 |
| Guides on e-cig use | 2.3% (0.0, 6.0) | 4.8% (0.0, 9.8) | 0.5% (0.0, 1.3) | 0.44 | 0.10 | 0.34 |
| E-cig product reviews | 12.6% (6.3, 18.9) | 33.2% (19.8, 46.5) | 16.2% (10.4, 22.1) | 0.01 | 0.02 | 0.40 |
| Lifestyle content involving e-cigs | 15.8% (8.0, 23.5) | 10.3% (0.0, 22.1) | 15.0% (9.4, 20.5) | 0.45 | 0.48 | 0.87 |
| Other | 2.2% (0.0, 5.3) | 1.7% (0.0, 5.0) | 0.1% (0.0, 0.4) | 0.84 | 0.36 | 0.20 |
| **N** | **150** | **61** | **239** |  |  |  |

**Table 3b:** Characteristics of E-Cigarette-Related Social Media Exposure (*continued*)

| **(1) Over-45 Exclusive Smokers; (2) Over-45 Exclusive Vapers; (3) Under-36 Smokers** |  |  |  |  |  |  |
| --- | --- | --- | --- | --- | --- | --- |
| Survey Measure | Weighted Prevalence (95% CI) |  |  | *p*-Value for Group Differences |  |  |
|  | (1) | (2) | (3) | (1) vs. (2) | (2) vs. (3) | (1) vs. (3) |
| *Platforms where content was seen* |  |  |  |  |  |  |
| Facebook | 45.6% (35.6, 55.6) | 49.4% (34.7, 64.0) | 57.6% (50.2, 65.1) | 0.67 | 0.33 | 0.06 |
| Instagram | 19.1% (11.2, 27.0) | 17.2% (7.4, 27.1) | 42.0% (34.5, 49.4) | 0.77 | 0.00 | 0.00 |
| TikTok | 27.9% (19.0, 36.8) | 24.0% (10.5, 37.4) | 46.1% (38.6, 53.5) | 0.63 | 0.00 | 0.00 |
| X/Twitter | 17.4% (9.5, 25.3) | 25.9% (12.1, 39.7) | 32.1% (25.2, 39.1) | 0.29 | 0.43 | 0.01 |
| YouTube | 52.7% (42.6, 62.7) | 46.1% (31.8, 60.5) | 53.4% (45.9, 60.9) | 0.46 | 0.38 | 0.91 |
| Other | 5.6% (0.7, 10.6) | 7.5% (0.3, 14.8) | 4.1% (1.2, 7.0) | 0.67 | 0.39 | 0.61 |
| **N** | **148** | **61** | **244** |  |  |  |
| *Content searched for or recommended* |  |  |  |  |  |  |
| No recs., all searches | 9.9% (4.0, 15.9) | 17.8% (7.3, 28.3) | 3.3% (0.4, 6.1) | 0.20 | 0.01 | 0.05 |
| Some recs., mostly searches | 6.9% (1.6, 12.1) | 20.2% (9.0, 31.4) | 4.1% (0.9, 7.4) | 0.03 | 0.01 | 0.38 |
| Mix of recs. and searches | 10.1% (4.6, 15.7) | 16.8% (6.3, 27.4) | 16.7% (11.1, 22.4) | 0.27 | 0.99 | 0.10 |
| Mostly recs., some searches | 11.1% (4.5, 17.6) | 5.0% (0.3, 9.8) | 12.4% (7.7, 17.1) | 0.15 | 0.03 | 0.75 |
| All recs., no searches | 62.0% (52.2, 71.8) | 40.1% (24.9, 55.3) | 63.5% (56.2, 70.8) | 0.02 | 0.01 | 0.81 |
| **N** | **146** | **59** | **237** |  |  |  |
| *How recently content was seen* |  |  |  |  |  |  |
| Within past week | 10.6% (5.1, 16.2) | 10.3% (3.2, 17.5) | 31.0% (23.8, 38.1) | 0.95 | 0.00 | 0.00 |
| Within past month | 28.9% (19.9, 37.9) | 28.7% (16.3, 41.0) | 36.9% (29.6, 44.3) | 0.98 | 0.26 | 0.17 |
| Within past 6 months | 31.6% (21.9, 41.4) | 34.7% (19.5, 49.8) | 20.0% (13.7, 26.3) | 0.74 | 0.08 | 0.05 |
| Longer than 6 months ago | 28.7% (19.5, 37.8) | 26.3% (14.6, 38.0) | 12.1% (7.0, 17.2) | 0.76 | 0.03 | 0.00 |
| **N** | **149** | **62** | **235** |  |  |  |
| *How many times content was seen* |  |  |  |  |  |  |
| Less than 10 | 41.5% (31.6, 51.5) | 31.0% (18.4, 43.6) | 30.0% (22.8, 37.2) | 0.20 | 0.89 | 0.07 |
| 10 to 20 | 32.6% (22.8, 42.3) | 33.5% (18.8, 48.1) | 33.6% (26.3, 40.9) | 0.92 | 0.99 | 0.87 |
| 20 to 50 | 17.1% (9.5, 24.6) | 22.8% (11.9, 33.7) | 20.3% (14.2, 26.5) | 0.40 | 0.70 | 0.51 |
| More than 50 | 8.8% (3.9, 13.8) | 12.7% (3.5, 22.0) | 16.0% (10.2, 21.8) | 0.47 | 0.55 | 0.06 |
| **N** | **148** | **63** | **230** |  |  |  |

*Notes:* Item response prevalences are post-stratification-weighted within each sampling group to match the 2023 NHIS by sex, race/ethnicity, and educational attainment (see Section 2.3.1 for details). *p*-values for differences in weighted responses across sampling groups are from design-adjusted Wald tests. 95% confidence intervals are shown in parentheses, with lower bounds truncated at zero when necessary. See Section 2.2.2 for exact definitions of the “derived any-topic” measures.

Regarding all topics of e-cigarette-related content viewed, the three most frequently indicated among over-45 exclusive smokers were health risks (64.4%), youth vaping (53.9%), and addiction (42.2%); for over-45 exclusive vapers, they were product reviews (66.0%), health risks (58.7%), and youth vaping (29.7%); and for under-36 smokers, they were health risks (63.9%), youth vaping (55.0%), and addiction (49.7%). The topic most frequently reported as “noticed most” was health risks among both older smokers (35.0%) and younger smokers (35.4%), while product reviews were most frequent among older vapers (32.2%). Compared with older vapers, older smokers were significantly more likely to indicate exposure to content about youth vaping (*p < .*01), but significantly less likely to indicate exposure to industry news (*p < .*05), product reviews (*p < .*01), and guides on e-cigarette use (*p < .*05). Compared with younger smokers, older smokers were significantly less likely to report exposure to industry news (*p < .*05) and product reviews (*p < .*05). These per-topic differences in exposure were reflected in the persuasive valence measures (whose construction is described in Section 2.2.2): older smokers were significantly more likely than both older vapers (*p < .*01) and younger smokers (*p < .*05) to report exposure to only negative (e.g., anti-vaping) topics. On average, the number of topics indicated was 2.6 for older smokers, 2.9 for older vapers, and 3.0 for younger smokers.

When asked whether e-cigarette–related content was searched for or recommended, the modal response for all groups was “all the content was recommended to me, I did not search for it,” comprising 62.0% of over-45 exclusive smokers, 40.1% of over-45 exclusive vapers, and 63.5% of under-36 smokers. Despite this shared modal tendency, both older smokers (*p < .*05) and younger smokers (*p < .*01) were significantly less likely than older vapers to indicate that all e-cigarette– related content was recommended. With respect to the recency of exposure, the majority of respondents in all three groups reported last viewing e-cigarette–related content more than a week prior. The modal response among older smokers (31.6%) and older vapers (34.7%) was “within the past 6 months,” while for younger smokers it was “within the past month” (36.9%). However, older smokers (10.6%) and older vapers (10.3%) were significantly less likely than younger smokers (31.0%) to report past-week exposure (*p < .*01 for both comparisons). In terms of cumulative exposure, a majority of each sampling group—older smokers (74.1%), older vapers (64.5%), and younger smokers (63.6%)—reported 20 or fewer instances of lifetime exposure to e-cigarette– related content.

Table 4 presents by-group associations between exposure to e-cigarette-related social media content and various attitudes toward e-cigarettes and vaping. Among over-45 exclusive smokers, exposure was positively associated with intentions for future e-cigarette use across multiple outcomes. In particular, exposure was significantly associated with reporting interest (vs. no interest) in using e-cigarettes in the future (OR=1.87, *p < .*01) and with reporting being likely (vs. unlikely or neutral) to consider switching to e-cigarettes if cigarette prices were to increase by $4 (OR=1.68, *p < .*05). Exposure was also positively associated with reporting that one would be likely (vs. unlikely or neutral) to consider switching if e-cigarettes were free (OR=1.37), though this estimate did not reach conventional significance levels (*p* = .13). Associations between exposure and these three outcomes remained positive across all alternative specifications (see Appendix D).

**Table 4:** Associations of E-Cigarette-Related Social Media Exposure with E-Cigarette Beliefs and Intentions (Logistic Regression)

| (1) Over-45 Exclusive Smokers; (2) Over-45 Exclusive Vapers; (3) Under-36 Smokers |  |  |  |  |  |  |
| --- | --- | --- | --- | --- | --- | --- |
| Outcome | Association with E-Cig Social Media Exposure, OR (95% CI) |  |  | N |  |  |
|  | (1) | (2) | (3) | (1) | (2) | (3) |
| **Intentions for future e-cig use** |  |  |  |  |  |  |
| *Interested in using e-cigs in future* | 1.87*** | — | — | **461** | — | — |
| (1=interested; 0=not interested) | (1.24, 2.81) | — | — |  |  |  |
| *How likely to consider switching if cigs increase \$4/pack* | 1.68** | — | — | **472** | — | — |
| (1=likely; 0=neutral/unlikely) | (1.12, 2.51) | — | — |  |  |  |
| *How likely to consider switching if e-cigs free* | 1.37 | — | — | **476** | — | — |
| (1=likely; 0=neutral/unlikely) | (0.91, 2.07) | — | — |  |  |  |
| **E-cig harm perceptions** |  |  |  |  |  |  |
| *How harmful are e-cigs for health* | 0.96 | 1.13 | 1.01 | **472** | **144** | **338** |
| (1=very/moderately harmful; 0=not at all/a little harmful) | (0.56, 1.66) | (0.57, 2.26) | (0.60, 1.71) |  |  |  |
| *Compared to cigs, how harmful are e-cigs for health* | 0.87 | 1.82 | 0.84 | **470** | **148** | **339** |
| (1=as/more harmful; 0=less harmful) | (0.56, 1.36) | (0.62, 5.34) | (0.51, 1.38) |  |  |  |
| *Compared to cigs, how addictive are e-cigs* | 0.70 | 0.99 | 1.11 | **457** | **148** | **338** |
| (1=as/more addictive; 0=less addictive) | (0.38, 1.27) | (0.44, 2.23) | (0.54, 2.24) |  |  |  |
| **Other e-cig attitudes** |  |  |  |  |  |  |
| *Gov. should spend to discourage youth vaping* | 1.50* | 1.50 | 0.91 | **471** | **149** | **337** |
| (1=agree; 0=neutral/disagree) | (1.00, 2.25) | (0.77, 2.95) | (0.56, 1.47) |  |  |  |
| *E-cigs are designed for adult smokers* | 1.43* | 2.10** | 1.00 | **459** | **149** | **334** |
| (1=agree; 0=neutral/disagree) | (0.94, 2.17) | (1.08, 4.08) | (0.61, 1.63) |  |  |  |
| *Many e-cig sales today are illicit* | 0.68 | 2.59* | 0.92 | **382** | **135** | **323** |
| (1=agree; 0=neutral/disagree) | (0.36, 1.30) | (0.98, 6.87) | (0.53, 1.61) |  |  |  |

Notes: \*\*\* $p < .01$ , \*\* $p < .05$ , \* $p < .1$ . Associations are estimated with binary logistic regression and reported as odds ratios (ORs) with 95% confidence intervals in parentheses. All estimates retain participants who opted out of the content exposure question, treating them the same as respondents who reported no exposure (coded 0). Outcomes on intentions for future e-cigarette use were asked only of over-45 exclusive smokers; all other outcomes were estimated on the full analytic sample. Participants who opted out of specific outcome questions were excluded from those models. Dichotomization cutoffs for each outcome are reported under the measure. All regressions controlled for sex, education, and race/ethnicity. Standard errors were estimated with Huber–White robust estimators, and significance was assessed using Wald tests.

Turning to perceived e-cigarette harms, assessed across all three sampling groups, exposure was not significantly associated with any of the three harm-perception outcomes. Among over-45 exclusive smokers, exposure was negatively associated with general perceptions of e-cigarette health risks (OR=0.96), with perceiving e-cigarettes as more or equivalently risky to health as cigarettes (OR=0.87), and with perceiving e-cigarettes as more or equivalently addictive as cigarettes (OR=0.70). However, all three associations were statistically insignificant, and the direction of the first two associations was sensitive to alternative specifications (see Appendix D). Among over-45 exclusive vapers, exposure was positively associated with perceptions of general (OR=1.13) and relative (OR=1.82) health risks—both insignificant but directionally robust. Finally, among under-36 smokers, exposure was negatively associated with relative health perceptions (OR=0.84) but positively associated with relative addictiveness perceptions (OR=1.11).

Exposure was positively associated with support for government spending to discourage youth vaping among both over-45 exclusive smokers (OR=1.50) and over-45 exclusive vapers (OR=1.50), with the former association significant at the 90% confidence level. Exposure was also significantly and positively associated with endorsement of the view that e-cigarettes are made “with adult smokers in mind” among both older smokers (OR=1.43, *p < .*1) and older vapers (OR=2.10, *p < .*05). Finally, exposure was negatively associated with agreement that “a lot of e-cigarette sales today are illicit” among older smokers (OR=0.68) and younger smokers (OR=0.92), though both associations were insignificant. In contrast, exposure was positively associated with agreement to this belief among older vapers (OR=2.59), reaching marginal significance (*p < .*1).

## 4 Discussion

This study examined the prevalence, nature, and attitudinal links of e-cigarette-related social media exposure across generations of nicotine users, with particular attention to how such exposure may relate to older smokers’ reluctance to adopt e-cigarettes as a reduced-harm alternative to cigarettes. Against this backdrop, two descriptive patterns stand out. First, older vapers were directionally more likely than older smokers to report exposure to e-cigarette content. Second, among older smokers, those exposed to such content were more likely to express interest in future e-cigarette use. In a setting with ideal identification, these are precisely the kinds of relationships one would expect if exposure plays a causal role in promoting e-cigarette adoption among older smokers.

However, non-causal explanations of these patterns remain plausible, especially given the empirical context. Selective attention induced by informational overload [48, 49], as well as engagement-optimized algorithmic targeting [50], suggest that those who already use e-cigarettes—or are inclined to use them in the future—would be more likely to report having seen e-cigarette-related content on online platforms. Furthermore, cross-group comparisons regarding whether content was recommended or searched for—in which older smokers were significantly more likely than older vapers to report encountering content exclusively through recommendations rather than searches— highlight the role for self-selection into exposure. These realities are consistent with a narrative in which the preference strength for e-cigarettes drives varying degrees of exposure: those with the strongest preferences for e-cigarettes (current vapers) deliberately seek content out; those with moderate preferences (smokers intending to vape) pay attention to recommended content; and those with the weakest preferences (smokers not intending to vape) either ignore such content or are not recommended it at all.

Triangulation with existing literature provides limited guidance in adjudicating whether the associations observed in this study reflect a causal effect of e-cigarette-related content exposure on e-cigarette adoption among older smokers. While surveys of younger populations often report similarly positive associations between exposure and stated intentions to vape [15, 16], the causal validity of these findings is subject to the same empirical ambiguities previously outlined. A cautious interpretation is especially warranted in light of experimental studies with younger samples, which collectively suggest that the effects of e-cigarette-related social media exposure are content-contingent: some types of content have been shown to encourage vaping intentions and behaviors [17, 18, 21, 22], while others appear to discourage use or have no effect [19–21]. At the same time, the significant age gradient in exposure (35.3% for older smokers vs. 72.0% for younger smokers) observed here parallels the age gradient in vaping rates from national data [28, 40], a directional consistency suggestive of a causal role for exposure. However, this pattern is also compatible with an alternative explanation wherein exposure exerts a positive effect only among younger users, which is plausible given age-dependent differences in factors such as cultural-historical associations with nicotine and tobacco products [51, 52] and long-run addiction profiles.

Nevertheless, if the exposure-promotes-vaping effect hinted at by this study’s findings does in fact apply to older smokers, additional results belie its operation through *semantic influence* or the promotion of more favorable objective beliefs about e-cigarettes and vaping. Among older smokers who reported exposure, *health risks of e-cigarettes* was the most frequently encountered topic (64.4%) and also the most commonly selected as the topic noticed most often (35.0%). In contrast, only 14.9% reported seeing content about the *health benefits of e-cigarettes for smokers*, and just 1.3% selected this as the topic noticed most often. Across all topics, older smokers were significantly more likely to report exposure to only negatively valenced (45.5%) than only positively valenced (19.8%) content (adjusted Wald test: *p < .*01). The overall pattern of these findings is corroborated by the associational analysis: exposure was not significantly associated with lower perceived e-cigarette harm on any of the three measures considered, and across multiple sensitivity specifications (see Tables D.1–D.3), exposure covaried positively with perceived harm. If exposure primarily promoted e-cigarette use through a semantic influence mechanism, one would expect exposed smokers to encounter more positively valenced content and to form more favorable beliefs about e-cigarette health risks as a result.

Although the results of this study run contrary to a narrative in which exposure promotes older-smoker e-cigarette uptake through favorable belief updating, they do not rule out the possibility of exposure promoting vaping via alternative mechanisms. In particular, exposure may promote vaping through a *mere exposure* mechanism, in which repeated visual exposure to e-cigarettes via social media increases the likelihood of use through heightened familiarity or affective resonance, without requiring any belief updating [53]. While the data of this study are not suited to test this mechanism directly, it aligns with several strands of prior evidence. Absent any explicit information provision about e-cigarettes, the presence of e-cigarettes in Instagram posts [17, 18] and music videos [22] has been shown to increase stated intentions to vape in randomized trials conducted among younger samples. Likewise, quasi-experimental studies of traditional media advertising find that exposure can increase reported vaping even when advertisements contain no explicit cessation or health claims [54]. A possible reconciliation, therefore, is that *semantic influence* and *mere exposure* processes operate in opposing directions among older smokers: negative informational content discourages use, while the affective familiarity generated through repeated visual exposure promotes it. The net positive association between exposure and interest in future vaping observed here could thus reflect the latter mechanism swamping the former among this population.

While the descriptive nature of the data prohibits firm conclusions about whether e-cigarette-related social media exposure increases the likelihood that older smokers adopt e-cigarettes, the findings nevertheless suggest several promising directions for future experimental research. One fruitful avenue would be to directly test the distinction between the mere exposure and semantic influence mechanisms. This could be done by exposing older smoker participants to Instagram-style posts featuring e-cigarettes, randomly varying whether the posts contain only imagery or also include valenced messaging, such as a modified risk statement (positive) or a warning label (negative). If the additional information has little incremental effect on e-cigarette beliefs or vaping intentions, this would provide causal support for mere exposure mechanistically dominating semantic influence in explaining how social media exposure promotes vaping. Another direction for future experimental work could test whether symbolic cues embedded in social media content— such as age-coded imagery, taglines, or stylistic elements—modulate its persuasive effectiveness. Testing whether older adults respond differently to content tailored to their generational identity versus youth-oriented aesthetics could inform the design of more effective harm-reduction messaging for older smokers. Furthermore, if combined with empirical work mapping how user demographics align with content distributions across platforms, such experiments could help clarify whether symbolic alignment contributes to age-related differences in the behavioral effects of e-cigarette-related social media exposure.

## 5 Conclusion

To the best of the author’s knowledge, this study provides the first focused assessment of the ecigarette-related social media experiences of older smokers. Given both their elevated disease risk and the fact that e-cigarettes remain an underutilized harm reduction tool for this population, understanding the role social media currently plays—and could play in the future—in facilitating switching is of substantial relevance to public health. While the cross-sectional nature of the data precludes firm causal conclusions, the analysis provides suggestive evidence that social media exposure may support the transition from smoking to vaping, and that such influence operates through advertising-style nudging rather than through correcting misinformation about e-cigarette health risks. These findings point to the potential of harm-reduction policies that increase older smokers’ exposure to neutral or positively valenced e-cigarette content on digital platforms—for example, by permitting e-cigarette advertising or promoting the algorithmic delivery of modified-risk information. However, such policy prescriptions remain premature until future research more clearly establishes the causal effects of exposure and the mechanisms through which they operate. Moreover, any effort to support older-smoker harm reduction through social media must weigh potential tradeoffs, including risks to user privacy and the possibility of encouraging nicotine-product initiation among youth.

## Abbreviations

NHIS: National Health Interview Survey

## Declarations

## Acknowledgements

The author thanks Asena Carner, Nicholas Grandstaff, Donald Kenkel, Alan Mathios, Jacob Meyer, and Hua Wang for helpful comments.

## Authors and Affiliations

Department of Economics, College of Arts and Sciences, Cornell University, Ithaca, New York, United States Ryan Dycus

## Author Contributions

R.D. was responsible for all aspects of the research and manuscript preparation, including study design, data collection, analysis, and drafting.

## Data Availability

A full replication package including all analyzed survey data is available from OSF (https://osf.io/dh9t2).

## Ethics Approval and Consent to Participate

Participants were fully informed about the study and its purpose, the voluntary nature of participation, and their right to withdraw at any time. All participants provided their express consent before starting the survey, which is recorded in the data. This study (protocol number IRB0149194) received exempt-status approval from Cornell’s IRB on November 7, 2024.

## Consent for Publication

Not applicable.

## Appendix A: Exact Wording of Survey Measures

### General Social Media Use

Do you regularly use social media? *Social media includes platforms such as YouTube, Instagram, Facebook, X/Twitter, and TikTok*.

- Yes
- No
- I’m unsure

Which social media platforms do you regularly use? *Please select all that apply*.

- Facebook
- Instagram
- TikTok
- X/Twitter
- YouTube
- Other (please specify)
- I don’t recall

How often do you use social media?

- Multiple times a day
- About once a day
- About once every few days
- About once a week
- About once every couple of weeks
- About once a month
- Other (please specify)

### E-Cigarette-Related Social Media Exposure

Have you ever viewed any content (posts, pictures, videos, memes, etc.) on social media discussing, depicting, or mentioning e-cigarettes or their use? *Social media includes platforms such as YouTube, Instagram, Facebook, X/Twitter, and TikTok*.

- Yes
- No
- I’m unsure

On which of the following social media platforms have you viewed content about e-cigarettes? *Please select all that apply*.

- Facebook
- Instagram
- TikTok
- X/Twitter
- YouTube
- Other (please specify)
- I don’t recall

When you’ve seen social media content about e-cigarettes or their use, which topics have you noticed? *Please select all that apply*.

- Health risks of e-cigarettes
- Health benefits of e-cigarettes for smokers
- E-cigarette industry news
- Youth e-cigarette use
- E-cigarette addiction
- Guides on how to use e-cigarettes
- Reviews of e-cigarette products
- Lifestyle content portraying e-cigarettes as cool, sexy, or fashionable
- Other (please specify)
- I don’t recall

When you’ve seen social media content about e-cigarettes or their use, which one topic have you noticed the most? *Please make a **single** selection*.

- Health risks of e-cigarettes
- Health benefits of e-cigarettes for smokers
- E-cigarette industry news
- Youth e-cigarette use
- E-cigarette addiction
- Guides on how to use e-cigarettes
- Reviews of e-cigarette products
- Lifestyle content portraying e-cigarettes as cool, sexy, or fashionable
- Other (please specify)
- I don’t recall

Was the content about e-cigarettes you viewed on social media recommended to you, or did you search for it yourself?

- All of the content was recommended to me, I did not search for it
- Most of the content was recommended to me, but some I searched for
- The content I viewed was a mix of recommendations and my own searches
- Some of the content was recommended to me, but mostly I searched for it
- None of the content was recommended to me, I searched for all of it
- I’m unsure

Considering posts, videos, pictures, and memes, about how many times have you ever seen ecigarette related content on social media?

- Less than 10 times
- 10 to 20 times
- 20 to 50 times
- 50 to 100 times
- 100 to 200 times
- More than 200 times
- I’m unsure

How recently have you viewed e-cigarette related content on social media?

- Within the past week
- Within the past month
- Within the past 6 months
- Within the past year
- Within the past 2 years
- More than 2 years ago
- I’m unsure

### Intentions for Future E-Cigarette Use

How interested are you in using e-cigarettes in the future?

- Not interested at all
- Somewhat interested
- Moderately interested
- Extremely interested
- I’m unsure

How likely is it that you would consider switching from smoking cigarettes to using e-cigarettes if the price you pay for cigarettes were to increase by $4 per pack?

- Extremely unlikely
- Somewhat unlikely
- Neither likely nor unlikely
- Somewhat likely
- Extremely likely
- I don’t know

How likely is it that you would consider switching from smoking cigarettes to using e-cigarettes if you did not have to pay for e-cigarettes?

- Extremely unlikely
- Somewhat unlikely
- Neither likely nor unlikely
- Somewhat likely
- Extremely likely
- I don’t know

### E-Cigarette Potential Harms

Overall, how harmful do you feel e-cigarettes are for a person’s health?

- Very harmful
- Moderately harmful
- A little harmful
- Not harmful at all
- I don’t know

Compared to smoking cigarettes, using e-cigarettes is…

- Much more harmful to health
- More *harmful to health*
- About as *harmful to health*
- Less *harmful to health*
- Much less *harmful to health*
- I don’t know

Compared to smoking cigarettes, using e-cigarettes is…

- Much more *addictive*
- More *addictive*
- About as *addictive*
- Less *addictive*
- Much less *addictive*
- I don’t know

### Other Attitudes about E-Cigarettes and Vaping

How much do you agree or disagree with the following statement? *The government should spend money on public service announcements aimed at discouraging youth away from using e-cigarettes*.

- Strongly agree
- Agree
- Neither agree nor disagree
- Disagree
- Strongly disagree
- I don’t know

How much do you agree or disagree with the following statement? *E-cigarettes are designed with adult smokers in mind*.

- Strongly agree
- Agree
- Neither agree nor disagree
- Disagree
- Strongly disagree
- I don’t know

How much do you agree or disagree with the following statement? *A lot of e-cigarette sales today are illicit or “under-the-table.”*

- Strongly agree
- Agree
- Neither agree nor disagree
- Disagree
- Strongly disagree
- I don’t know

## Appendix B: Unweighted Comparative Characterization Analysis

This appendix reproduces the comparative characterization analysis without post-stratification weighting. Raw prevalences of item responses are reported, and differences across sampling groups are assessed with two-sided Z-tests for differences in proportions. Table B.1 presents the unweighted analog of Table 2 (characteristics of general social media use) and Tables B.2a–b present the unweighted analogs of Tables 3a–b (characteristics of e-cigarette-related social media exposure).

**Table B.1:** (Unweighted) Characteristics of General Social Media Use.

| **(1) Over-45 Exclusive Smokers; (2) Over-45 Exclusive Vapers; (3) Under-36 Smokers** |  |  |  |  |  |  |
| --- | --- | --- | --- | --- | --- | --- |
| Survey Measure | Weighted Prevalence (95% CI) |  |  | *p*-Value for Group Differences |  |  |
|  | (1) | (2) | (3) | (1) vs. (2) | (2) vs. (3) | (1) vs. (3) |
| *Use Social Media Regularly?* |  |  |  |  |  |  |
| Yes | 91.9% | 94.0% | 95.0% | 0.38 | 0.66 | 0.08 |
| No | 8.1% | 6.0% | 5.0% | 0.38 | 0.66 | 0.08 |
| **N** | **483** | **149** | **338** |  |  |  |
| *Social Media Use Frequency* |  |  |  |  |  |  |
| Less often than daily | 5.4% | 7.9% | 1.9% | 0.33 | 0.01 | 0.01 |
| Once per day | 19.6% | 18.6% | 9.7% | 0.78 | 0.02 | 0.00 |
| Multiple times per day | 74.9% | 73.6% | 88.5% | 0.75 | 0.00 | 0.00 |
| **N** | **443** | **140** | **321** |  |  |  |
| *Platforms Used* |  |  |  |  |  |  |
| Facebook | 84.0% | 81.4% | 81.6% | 0.50 | 0.96 | 0.40 |
| Instagram | 48.3% | 49.3% | 72.0% | 0.84 | 0.00 | 0.00 |
| TikTok | 42.0% | 33.6% | 60.4% | 0.07 | 0.00 | 0.00 |
| X/Twitter | 33.6% | 43.6% | 57.3% | 0.04 | 0.01 | 0.00 |
| YouTube | 77.2% | 78.6% | 84.4% | 0.73 | 0.15 | 0.01 |
| Other | 10.2% | 10.7% | 6.9% | 0.85 | 0.20 | 0.10 |
| **N** | **443** | **140** | **321** |  |  |  |

*Notes:* *p*-values reflect two-sided Z-tests for differences in proportions across sampling groups.

**Table B.2a:** (Unweighted) Characteristics of E-Cigarette-Related Social Media Exposure.

| **(1) Over-45 Exclusive Smokers; (2) Over-45 Exclusive Vapers; (3) Under-36 Smokers** |  |  |  |  |  |  |
| --- | --- | --- | --- | --- | --- | --- |
| Survey Measure | Weighted Prevalence (95% CI) |  |  | *p*-Value for Group Differences |  |  |
|  | (1) | (2) | (3) | (1) vs. (2) | (2) vs. (3) | (1) vs. (3) |
| *Ever seen e-cig social media content* |  |  |  |  |  |  |
| Yes | 32.6% | 43.0% | 72.7% | 0.03 | 0.00 | 0.00 |
| No | 52.1% | 47.0% | 22.0% | 0.28 | 0.00 | 0.00 |
| Don't recall | 15.3% | 10.1% | 5.3% | 0.08 | 0.08 | 0.00 |
| **N** | **484** | **149** | **341** |  |  |  |
| *Any topics of content noticed* |  |  |  |  |  |  |
| E-cig health risks | 60.8% | 57.1% | 64.9% | 0.62 | 0.27 | 0.41 |
| E-cig health benefits for smokers | 17.0% | 25.4% | 19.0% | 0.18 | 0.29 | 0.61 |
| Youth vaping | 52.9% | 30.2% | 55.0% | 0.00 | 0.00 | 0.70 |
| E-cig industry news | 15.0% | 31.7% | 27.3% | 0.01 | 0.50 | 0.00 |
| E-cig addiction | 42.5% | 28.6% | 51.2% | 0.05 | 0.00 | 0.09 |
| Guides on e-cig use | 13.1% | 25.4% | 16.5% | 0.05 | 0.14 | 0.34 |
| E-cig product reviews | 30.1% | 63.5% | 40.5% | 0.00 | 0.00 | 0.03 |
| Lifestyle content involving e-cigs | 30.7% | 17.5% | 34.7% | 0.03 | 0.00 | 0.41 |
| Other | 4.6% | 7.9% | 1.7% | 0.38 | 0.08 | 0.12 |
| Num. of topics indicated (mean) | 2.7 | 2.9 | 3.1 | 0.32 | 0.23 | 0.00 |
| **N** | **153** | **63** | **242** |  |  |  |
| *Derived topical valence measures* |  |  |  |  |  |  |
| Only positive topics seen | 20.5% | 27.9% | 11.7% | 0.27 | 0.01 | 0.02 |
| Only negative topics seen | 41.7% | 24.6% | 32.5% | 0.01 | 0.21 | 0.07 |
| Both positive and negative topics seen | 11.8% | 19.5% | 39.3% | 0.03 | 0.00 | 0.00 |
| **N** | **151** | **61** | **240** |  |  |  |
| *Topic of content noticed most* |  |  |  |  |  |  |
| E-cig health risks | 36.0% | 29.5% | 38.5% | 0.36 | 0.18 | 0.62 |
| E-cig health benefits for smokers | 2.0% | 3.3% | 3.8% | 0.62 | 0.85 | 0.30 |
| Youth vaping | 20.0% | 8.2% | 14.2% | 0.01 | 0.15 | 0.15 |
| E-cig industry news | 2.0% | 8.2% | 6.7% | 0.10 | 0.70 | 0.02 |
| E-cig addiction | 8.0% | 4.9% | 9.6% | 0.39 | 0.17 | 0.58 |
| Guides on e-cig use | 1.3% | 6.6% | 0.8% | 0.12 | 0.08 | 0.65 |
| E-cig product reviews | 14.0% | 31.1% | 11.7% | 0.01 | 0.00 | 0.52 |
| Lifestyle content involving e-cigs | 14.7% | 6.6% | 14.2% | 0.06 | 0.05 | 0.90 |
| Other | 2.0% | 1.6% | 0.4% | 0.86 | 0.47 | 0.20 |
| **N** | **150** | **61** | **239** |  |  |  |

**Table B.2b:** (Unweighted) Characteristics of E-Cigarette-Related Social Media Exposure (*continuted*)

| **(1) Over-45 Exclusive Smokers; (2) Over-45 Exclusive Vapers; (3) Under-36 Smokers** |  |  |  |  |  |  |
| --- | --- | --- | --- | --- | --- | --- |
| Survey Measure | Weighted Prevalence (95% CI) |  |  | *p*-Value for Group Differences |  |  |
|  | (1) | (2) | (3) | (1) vs. (2) | (2) vs. (3) | (1) vs. (3) |
| *Platforms where content was seen* |  |  |  |  |  |  |
| Facebook | 52.7% | 47.5% | 58.6% | 0.50 | 0.12 | 0.26 |
| Instagram | 19.6% | 19.7% | 44.7% | 0.99 | 0.00 | 0.00 |
| TikTok | 29.1% | 21.3% | 51.2% | 0.23 | 0.00 | 0.00 |
| X/Twitter | 16.9% | 23.0% | 38.5% | 0.33 | 0.01 | 0.00 |
| YouTube | 45.9% | 52.5% | 49.6% | 0.39 | 0.69 | 0.48 |
| Other | 4.7% | 6.6% | 4.5% | 0.62 | 0.55 | 0.92 |
| **N** | **148** | **61** | **244** |  |  |  |
| *Content searched for or recommended* |  |  |  |  |  |  |
| No recs., all searches | 10.3% | 16.9% | 2.5% | 0.23 | 0.00 | 0.00 |
| Some recs., mostly searches | 6.8% | 20.3% | 3.0% | 0.02 | 0.00 | 0.10 |
| Mix of recs. and searches | 11.0% | 15.3% | 18.1% | 0.43 | 0.59 | 0.05 |
| Mostly recs., some searches | 8.9% | 8.5% | 19.4% | 0.92 | 0.01 | 0.00 |
| All recs., no searches | 63.0% | 39.0% | 57.0% | 0.00 | 0.01 | 0.24 |
| **N** | **146** | **59** | **237** |  |  |  |
| *How recently content was seen* |  |  |  |  |  |  |
| Within past week | 12.1% | 14.5% | 32.3% | 0.64 | 0.00 | 0.00 |
| Within past month | 30.2% | 29.0% | 40.4% | 0.87 | 0.09 | 0.04 |
| Within past 6 months | 29.5% | 27.4% | 17.0% | 0.76 | 0.10 | 0.01 |
| Longer than 6 months ago | 27.5% | 29.0% | 10.2% | 0.83 | 0.00 | 0.00 |
| **N** | **149** | **62** | **235** |  |  |  |
| *How many times content was seen* |  |  |  |  |  |  |
| Less than 10 | 41.9% | 31.7% | 29.1% | 0.16 | 0.69 | 0.01 |
| 10 to 20 | 29.1% | 31.7% | 35.2% | 0.70 | 0.60 | 0.21 |
| 20 to 50 | 18.2% | 25.4% | 19.1% | 0.26 | 0.31 | 0.83 |
| More than 50 | 10.8% | 11.1% | 16.5% | 0.95 | 0.25 | 0.11 |
| **N** | **148** | **63** | **230** |  |  |  |

*Notes:* *p*-values reflect two-sided Z-tests for differences in proportions across sampling groups. See Section 2.2.2 for exact definitions of the “derived any-topic” measures.

## Appendix C: Summary of Responses to Associational Analysis Outcome Measures

This appendix provides by-group summaries of responses to the outcome measures used in the associational analysis across the same dichotomous cutoffs used in the main analysis (Table 4). Table C.1 summarizes responses to questions regarding participants’ intentions for future e-cigarette use, which were asked only of over-45 exclusive smokers. Table C.2 summarizes all other outcome measures by sampling group. Both tables report weighted prevalences of item responses, using the same post-stratification weighting procedure as in the comparative characterization analysis of the social media questions (within-group weighting to match the 2023 NHIS by sex, educational attainment, and race/ethnicity). Table C.2 also compares responses across sampling groups, reporting *p*-values from design-adjusted Wald tests.

**Table C.1:**
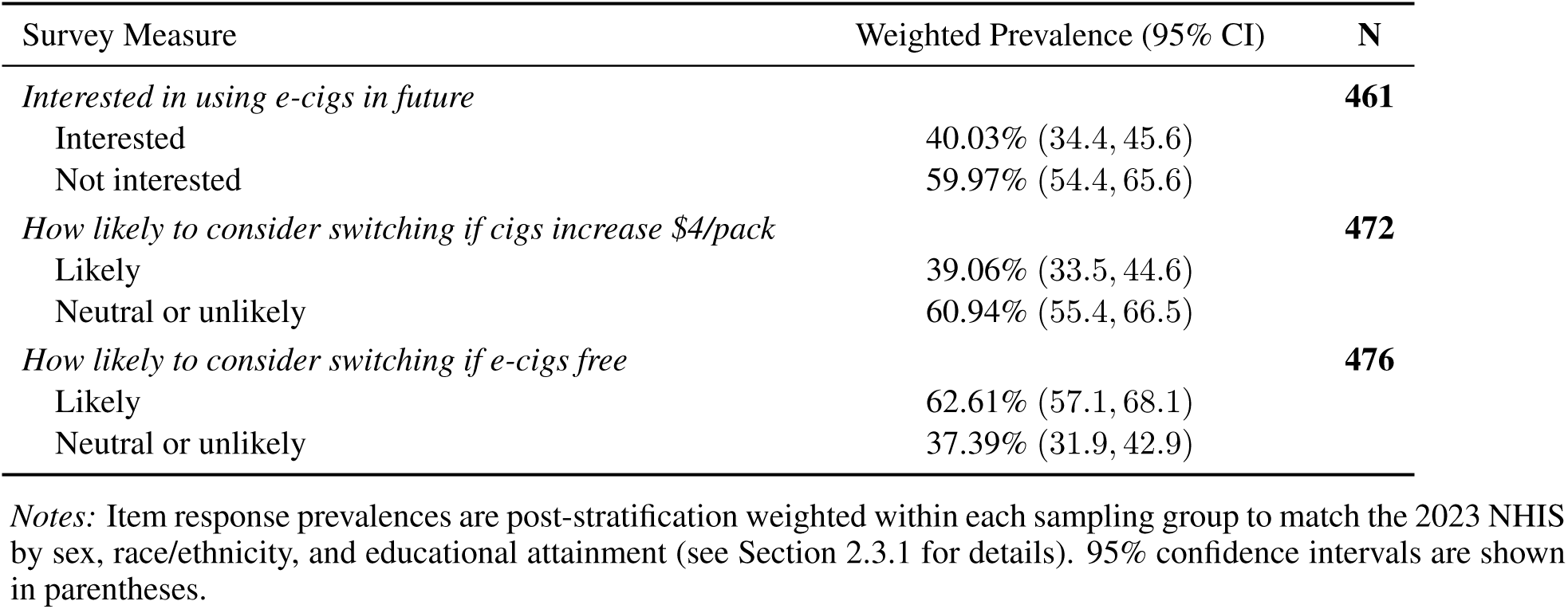
Summary of Responses to Intentions for Future E-Cigarette Use Questions (Over-45 Exclusive Smokers Only)

**Table C.2:** Summary of Responses to Harm Perception and Other E-Cigarette Attitude Questions.

| (1) Over-45 Exclusive Smokers; (2) Over-45 Exclusive Vapers; (3) Under-36 Smokers |  |  |  |  |  |  |
| --- | --- | --- | --- | --- | --- | --- |
| Survey Measure | Weighted Prevalence (95% CI) |  |  | *p*-Value for Group Differences |  |  |
|  | (1) | (2) | (3) | (1) vs. (2) | (2) vs. (3) | (1) vs. (3) |
| *How harmful are e-cigs for health* |  |  |  |  |  |  |
| Very or moderately harmful | 85.4% (81.4, 89.4) | 39.1% (30.1, 48.1) | 73.8% (68.3, 79.3) | 0.00 | 0.00 | 0.00 |
| A little or not at all harmful | 14.6% (10.6, 18.6) | 60.9% (51.9, 69.9) | 26.2% (20.7, 31.7) | 0.00 | 0.00 | 0.00 |
| **N** | **472** | **144** | **338** |  |  |  |
| *Compared to cigs, how harmful are e-cigs for health* |  |  |  |  |  |  |
| As or more harmful | 73.3% (68.1, 78.4) | 10.9% (4.6, 17.2) | 60.2% (54.0, 66.5) | 0.00 | 0.00 | 0.00 |
| Less harmful | 26.7% (21.6, 31.9) | 89.1% (82.8, 95.4) | 39.8% (33.5, 46.0) | 0.00 | 0.00 | 0.00 |
| **N** | **470** | **148** | **339** |  |  |  |
| *Compared to cigs, how addictive are e-cigs* |  |  |  |  |  |  |
| As or more addictive | 87.7% (83.8, 91.7) | 76.5% (68.5, 84.4) | 89.1% (85.3, 92.9) | 0.01 | 0.01 | 0.63 |
| Less addictive | 12.3% (8.3, 16.2) | 23.5% (15.6, 31.5) | 10.9% (7.1, 14.7) | 0.01 | 0.01 | 0.63 |
| **N** | **457** | **148** | **338** |  |  |  |
| *Gov. should spend to discourage youth vaping* |  |  |  |  |  |  |
| Agree | 59.2% (53.5, 64.8) | 46.1% (37.1, 55.2) | 52.8% (46.5, 59.2) | 0.02 | 0.23 | 0.15 |
| Neutral or disagree | 40.8% (35.2, 46.5) | 53.9% (44.8, 62.9) | 47.2% (40.8, 53.5) | 0.02 | 0.23 | 0.15 |
| **N** | **471** | **149** | **337** |  |  |  |
| *E-cigs are designed for adult smokers* |  |  |  |  |  |  |
| Agree | 29.5% (24.3, 34.8) | 49.9% (40.7, 59.0) | 42.8% (36.5, 49.1) | 0.00 | 0.21 | 0.00 |
| Neutral or disagree | 70.5% (65.2, 75.7) | 50.1% (41.0, 59.3) | 57.2% (50.9, 63.5) | 0.00 | 0.21 | 0.00 |
| **N** | **459** | **149** | **334** |  |  |  |
| *Many e-cig sales today are illicit* |  |  |  |  |  |  |
| Agree | 12.6% (8.7, 16.6) | 15.1% (7.9, 22.3) | 25.0% (19.4, 30.7) | 0.55 | 0.03 | 0.00 |
| Neutral or disagree | 87.4% (83.4, 91.3) | 84.9% (77.7, 92.1) | 75.0% (69.3, 80.6) | 0.55 | 0.03 | 0.00 |
| **N** | **382** | **135** | **323** |  |  |  |

*Notes:* Item response prevalences are post-stratification weighted within each sampling group to match the 2023 NHIS by sex, race/ethnicity, and educational attainment (see Section 2.3.1 for details). *p*-values for differences in weighted responses across sampling groups are from adjusted Wald tests. 95% confidence intervals are shown in parentheses, with lower bounds truncated at zero when necessary.

## Appendix D: Sensitivity Analysis

This appendix reports sensitivity analyses of the sampling-group–specific associations presented in Table 4. Table D.1 reproduces the binary logistic regressions from Table 4 but excludes respondents who opted out of the exposure question, rather than coding them as 0 as in the main analysis. Table D.2 presents ordinal logistic regression models, with all outcomes collapsed into three categories in a manner that balances concerns about overfitting with interpretability. Using these same three-category outcomes, Table D.3 reports binary logistic regressions estimated at both possible cutoffs for each measure. Finally, Table D.4 reproduces the primary binary logistic regression models using the post-stratification weights applied in the characterization analysis. Model specifications in Tables D.2 and D.3 are otherwise identical in structure to those in the main analysis.

**Table D.1:** Associations of E-Cigarette-Related Social Media Exposure with E-Cigarette Beliefs and Intentions (Logistic Regression, Opt-Outs Excluded)

| (1) Over-45 Exclusive Smokers; (2) Over-45 Exclusive Vapers; (3) Under-36 Smokers |  |  |  |  |  |  |
| --- | --- | --- | --- | --- | --- | --- |
| Outcome | Association with E-Cig Social Media Exposure, OR (95% CI) |  |  | N |  |  |
|  | (1) | (2) | (3) | (1) | (2) | (3) |
| **Intentions for future e-cig use** |  |  |  |  |  |  |
| *Interested in using e-cigs in future* | 1.60** | — | — | **391** | — | — |
| (1=interested; 0=not interested) | (1.05, 2.45) | — | — |  |  |  |
| *How likely to consider switching if cigs increase \$4/pack* | 1.48* | — | — | **400** | — | — |
| (1=likely; 0=neutral/unlikely) | (0.98, 2.25) | — | — |  |  |  |
| *How likely to consider switching if e-cigs free* | 1.23 | — | — | **405** | — | — |
| (1=likely; 0=neutral/unlikely) | (0.80, 1.88) | — | — |  |  |  |
| **E-cig harm perceptions** |  |  |  |  |  |  |
| *How harmful are e-cigs for health* | 1.10 | 1.12 | 1.03 | **401** | **131** | **321** |
| (1=very/moderately harmful; 0=not at all/a little harmful) | (0.63, 1.92) | (0.55, 2.31) | (0.58, 1.81) |  |  |  |
| *Compared to cigs, how harmful are e-cigs for health* | 1.03 | 1.68 | 0.89 | **397** | **133** | **321** |
| (1=as/more harmful; 0=less harmful) | (0.65, 1.62) | (0.55, 5.19) | (0.52, 1.52) |  |  |  |
| *Compared to cigs, how addictive are e-cigs* | 0.69 | 0.97 | 1.04 | **386** | **133** | **320** |
| (1=as/more addictive; 0=less addictive) | (0.36, 1.32) | (0.41, 2.26) | (0.48, 2.25) |  |  |  |
| **Other e-cig attitudes** |  |  |  |  |  |  |
| *Gov. should spend to discourage youth vaping* | 1.51* | 1.42 | 1.01 | **400** | **134** | **320** |
| (1=agree; 0=neutral/disagree) | (0.99, 2.31) | (0.71, 2.85) | (0.60, 1.69) |  |  |  |
| *E-cigs are designed for adult smokers* | 1.45* | 2.21** | 0.91 | **390** | **134** | **316** |
| (1=agree; 0=neutral/disagree) | (0.93, 2.24) | (1.10, 4.43) | (0.54, 1.55) |  |  |  |
| *Many e-cig sales today are illicit* | 0.60 | 2.54* | 0.80 | **326** | **122** | **306** |
| (1=agree; 0=neutral/disagree) | (0.31, 1.17) | (0.92, 7.07) | (0.44, 1.45) |  |  |  |

Notes: \*\*\* $p < .01$ , \*\* $p < .05$ , \* $p < .1$ . Associations are estimated with binary logistic regression and reported as odds ratios (ORs) with 95% confidence intervals in parentheses. All estimates drop participants who opted out of the content exposure question. Outcomes on intentions for future e-cigarette use were asked only of over-45 exclusive smokers; all other outcomes were estimated on the full analytic sample. Participants who opted out of specific outcome questions were excluded from those models. Dichotomization cutoffs for each outcome are reported under the measure. All regressions controlled for sex, education, and race/ethnicity. Standard errors were estimated with Huber–White robust estimators, and significance was assessed using Wald tests.

**Table D.2:** Associations of E-Cigarette-Related Social Media Exposure with E-Cigarette Beliefs and Intentions (Ordinal Logistic Regression)

| (1) Over-45 Exclusive Smokers; (2) Over-45 Exclusive Vapers; (3) Under-36 Smokers |  |  |  |  |  |  |
| --- | --- | --- | --- | --- | --- | --- |
| Outcome | Association with E-Cig Social Media Exposure, OR (95% CI) |  |  | N |  |  |
|  | (1) | (2) | (3) | (1) | (2) | (3) |
| **Intentions for future e-cig use** |  |  |  |  |  |  |
| *Interested in using e-cigs in future* | 1.82*** | — | — | **461** | — | — |
| (1=interested; 0=not interested) | (1.23, 2.69) | — | — |  |  |  |
| *How likely to consider switching if cigs increase \$4/pack* | 1.60** | — | — | **472** | — | — |
| (1=likely; 0=neutral/unlikely) | (1.09, 2.35) | — | — |  |  |  |
| *How likely to consider switching if e-cigs free* | 1.44* | — | — | **476** | — | — |
| (1=likely; 0=neutral/unlikely) | (1.00, 2.07) | — | — |  |  |  |
| **E-cig harm perceptions** |  |  |  |  |  |  |
| *How harmful are e-cigs for health* | 1.23 | 1.24 | 1.15 | **472** | **144** | **338** |
| (1=very/moderately harmful; 0=not at all/a little harmful) | (0.86, 1.76) | (0.62, 2.47) | (0.72, 1.84) |  |  |  |
| *Compared to cigs, how harmful are e-cigs for health* | 0.97 | 1.94 | 0.94 | **470** | **148** | **339** |
| (1=as/more harmful; 0=less harmful) | (0.67, 1.40) | (0.66, 5.69) | (0.60, 1.46) |  |  |  |
| *Compared to cigs, how addictive are e-cigs* | 0.83 | 0.96 | 1.51 | **457** | **148** | **338** |
| (1=as/more addictive; 0=less addictive) | (0.57, 1.19) | (0.48, 1.89) | (0.87, 2.63) |  |  |  |
| **Other e-cig attitudes** |  |  |  |  |  |  |
| *Gov. should spend to discourage youth vaping* | 1.58** | 1.30 | 0.96 | **471** | **149** | **337** |
| (1=agree; 0=neutral/disagree) | (1.07, 2.34) | (0.66, 2.57) | (0.61, 1.53) |  |  |  |
| *E-cigs are designed for adult smokers* | 1.12 | 1.90** | 0.99 | **459** | **149** | **334** |
| (1=agree; 0=neutral/disagree) | (0.77, 1.63) | (1.08, 3.35) | (0.62, 1.59) |  |  |  |
| *Many e-cig sales today are illicit* | 1.00 | 1.68 | 1.04 | **382** | **135** | **323** |
| (1=agree; 0=neutral/disagree) | (0.67, 1.50) | (0.82, 3.45) | (0.63, 1.71) |  |  |  |

Notes: \*\*\* $p < .01$ , \*\* $p < .05$ , \* $p < .1$ . Proportional-odds associations are estimated with ordinal logistic regression and reported as odds ratios (ORs) with 95% confidence intervals in parentheses. All estimates retain participants who opted out of the content exposure question, treating them the same as respondents who reported no exposure (coded 0). Outcomes on intentions for future e-cigarette use were asked only of over-45 exclusive smokers; all other outcomes were estimated on the full analytic sample. Participants who opted out of specific outcome questions were excluded from those models. Cutoffs for each outcome are reported under the measure. All regressions controlled for sex, education, and race/ethnicity. Standard errors were estimated with Huber–White robust estimators, and significance was assessed using Wald tests.

**Table D.3:** Associations of E-Cigarette-Related Social Media Exposure with E-Cigarette Beliefs and Intentions (Logistic Regression, Alternative Cutoffs)

| (1) Over-45 Exclusive Smokers; (2) Over-45 Exclusive Vapers; (3) Under-36 Smokers |  |  |  |  |  |  |
| --- | --- | --- | --- | --- | --- | --- |
| Outcome | Association with E-Cig Social Media Exposure, OR |  |  |  |  |  |
|  | Top-Category (=3) |  |  | Top-Two-Category (=2 or 3) |  |  |
|  | (1) | (2) | (3) | (1) | (2) | (3) |
| **Intentions for future e-cig use** |  |  |  |  |  |  |
| *Interested in using e-cigs in future* | 1.70* | — | — | 1.87*** | — | — |
| (1=not at all interested; 2=somewhat interested; 3=moderately/very interested) | (0.96, 3.01) | — | — | (1.24, 2.81) | — | — |
| *How likely to consider switching if cigs increase \$4/pack* | 1.37 | — | — | 1.68** | — | — |
| (1=neutral/unlikely; 2=somewhat likely; 3=extremely likely) | (0.76, 2.45) | — | — | (1.12, 2.51) | — | — |
| *How likely to consider switching if e-cigs free* | 1.54** | — | — | 1.37 | — | — |
| (1=neutral/unlikely; 2=somewhat likely; 3=extremely likely) | (1.00, 2.35) | — | — | (0.91, 2.07) | — | — |
| **Potential e-cig harm perceptions** |  |  |  |  |  |  |
| *How harmful are e-cigs for health* | 1.36 | 2.43 | 1.26 | 0.96 | 1.13 | 1.01 |
| (1=not at all/a little harmful; 2=moderately harmful; 3=very harmful) | (0.91, 2.03) | (0.76, 7.82) | (0.74, 2.14) | (0.56, 1.66) | (0.57, 2.26) | (0.60, 1.71) |
| *Compared to cigs, how harmful are e-cigs for health* | 1.08<sup>a</sup> | 10.13<sup>a</sup> | 1.22<sup>a</sup> | 0.87 | 1.82 | 0.84 |
| (1=less harmful; 2=as harmful; 3=more harmful) | (0.68, 1.71)<sup>a</sup> | (0.51, 200.09)<sup>a</sup> | (0.61, 2.42)<sup>a</sup> | (0.56, 1.36) | (0.62, 5.34) | (0.51, 1.38) |
| *Compared to cigs, how addictive are e-cigs* | 0.89 | 0.89 | 1.58* | 0.70 | 0.99 | 1.11 |
| (1=less addictive; 2=as addictive; 3=more addictive) | (0.53, 1.49) | (0.34, 2.33) | (0.93, 2.66) | (0.38, 1.27) | (0.44, 2.23) | (0.54, 2.24) |
| **Other e-cig attitudes** |  |  |  |  |  |  |
| *Gov. should spend to discourage youth vaping* | 1.50* | 1.50 | 0.91 | 1.85** | 1.00 | 1.07 |
| (1=disagree; 2=neutral; 3=agree) | (1.00, 2.25) | (0.77, 2.95) | (0.56, 1.47) | (1.03, 3.32) | (0.47, 2.12) | (0.60, 1.89) |
| *E-cigs are designed for adult smokers* | 1.43* | 2.10** | 1.00 | 0.92 | 1.77 | 1.01 |
| (1=disagree; 2=neutral; 3=agree) | (0.94, 2.17) | (1.08, 4.08) | (0.61, 1.63) | (0.62, 1.37) | (0.77, 4.11) | (0.61, 1.68) |
| *Many e-cig sales today are illicit* | 0.68 | 2.59* | 0.92 | 1.08 | 1.48 | 1.13 |
| (1=disagree; 2=neutral; 3=agree) | (0.36, 1.30) | (0.98, 6.87) | (0.53, 1.61) | (0.69, 1.67) | (0.71, 3.07) | (0.69, 1.85) |

Notes: \*\*\* $p < .01$ , \*\* $p < .05$ , \* $p < .1$ . Associations are estimated with binary logistic regression and reported as odds ratios (ORs) with 95% confidence intervals in parentheses. All estimates retain participants who opted out of the content exposure question, treating them the same as respondents who reported no exposure (coded 0). Outcomes on intentions for future e-cigarette use were asked only of over-45 exclusive smokers; all other outcomes were estimated on the full analytic sample. Participants who opted out of specific outcome questions were excluded from those models. Cutoffs for each outcome are reported under the measure. All regressions controlled for sex, education, and race/ethnicity. All results—except those denoted by <sup>a</sup>—were estimated with maximum likelihood with Huber-White robust standard errors. Estimates denoted by <sup>a</sup> were obtained via Firth's penalized maximum likelihood. See Table D.2 for by-group sample sizes.

**Table D.4:** Associations of E-Cigarette-Related Social Media Exposure with E-Cigarette Beliefs and Intentions (Logistic Regression, Post-Stratification Weighted)

| (1) Over-45 Exclusive Smokers; (2) Over-45 Exclusive Vapers; (3) Under-36 Smokers |  |  |  |  |  |  |
| --- | --- | --- | --- | --- | --- | --- |
| Outcome | Association with E-Cig Social Media Exposure, OR (95% CI) |  |  | N |  |  |
|  | (1) | (2) | (3) | (1) | (2) | (3) |
| **Intentions for future e-cig use** |  |  |  |  |  |  |
| *Interested in using e-cigs in future* | 1.56* | — | — | **461** | — | — |
| (1=interested; 0=not interested) | (0.93, 2.61) | — | — |  |  |  |
| *How likely to consider switching if cigs increase \$4/pack* | 1.62* | — | — | **472** | — | — |
| (1=likely; 0=neutral/unlikely) | (0.97, 2.69) | — | — |  |  |  |
| *How likely to consider switching if e-cigs free* | 1.04 | — | — | **476** | — | — |
| (1=likely; 0=neutral/unlikely) | (0.63, 1.73) | — | — |  |  |  |
| **E-cig harm perceptions** |  |  |  |  |  |  |
| *How harmful are e-cigs for health* | 1.06 | 1.31 | 1.35 | **472** | **144** | **338** |
| (1=very/moderately harmful; 0=not at all/a little harmful) | (0.54, 2.09) | (0.61, 2.78) | (0.72, 2.54) |  |  |  |
| *Compared to cigs, how harmful are e-cigs for health* | 0.84 | 2.16 | 0.81 | **470** | **148** | **339** |
| (1=as/more harmful; 0=less harmful) | (0.48, 1.45) | (0.66, 7.13) | (0.45, 1.46) |  |  |  |
| *Compared to cigs, how addictive are e-cigs* | 0.60 | 1.61 | 1.44 | **457** | **148** | **338** |
| (1=as/more addictive; 0=less addictive) | (0.28, 1.27) | (0.66, 3.93) | (0.61, 3.36) |  |  |  |
| **Other e-cig attitudes** |  |  |  |  |  |  |
| *Gov. should spend to discourage youth vaping* | 1.44 | 1.47 | 0.94 | **471** | **149** | **337** |
| (1=agree; 0=neutral/disagree) | (0.86, 2.42) | (0.70, 3.08) | (0.53, 1.65) |  |  |  |
| *E-cigs are designed for adult smokers* | 1.65* | 2.25** | 0.93 | **459** | **149** | **334** |
| (1=agree; 0=neutral/disagree) | (0.97, 2.78) | (1.08, 4.71) | (0.52, 1.66) |  |  |  |
| *Many e-cig sales today are illicit* | 0.68 | 3.03** | 1.00 | **382** | **135** | **323** |
| (1=agree; 0=neutral/disagree) | (0.31, 1.51) | (1.03, 8.91) | (0.51, 1.95) |  |  |  |

Notes: \*\*\* $p < .01$ , \*\* $p < .05$ , \* $p < .1$ . Associations are estimated with binary logistic regression and reported as odds ratios (ORs) with 95% confidence intervals in parentheses. All regressions apply post-stratification weights constructed within each sampling group to match the 2023 NHIS by sex, race/ethnicity, and educational attainment (see Section 2.3.1 for details). All estimates retain participants who opted out of the content exposure question, treating them the same as respondents who reported no exposure (coded 0). Outcomes on intentions for future e-cigarette use were asked only of over-45 exclusive smokers; all other outcomes were estimated on the full analytic sample. Participants who opted out of specific outcome questions were excluded from those models. Dichotomization cutoffs for each outcome are reported under the measure. All regressions controlled for sex, education, and race/ethnicity. Standard errors were estimated with Huber–White robust estimators, and significance was assessed using Wald tests.

